# Self-Focused Brain Predictors of Cognitive Behavioral Therapy Response in a Transdiagnostic Sample

**DOI:** 10.1101/2023.08.30.23294878

**Authors:** Angela Fang, Bengi Baran, Jamie D. Feusner, K. Luan Phan, Clare C. Beatty, Jessica Crane, Ryan J. Jacoby, Dara S. Manoach, Sabine Wilhelm

## Abstract

**Background:** Effective biomarkers of cognitive behavioral therapy (CBT) response provide information beyond available behavioral or self-report measures and may optimize treatment selection for patients based on likelihood of benefit. No single biomarker reliably predicts CBT response. In this study, we evaluated patterns of brain connectivity associated with self-focused attention (SFA) as biomarkers of CBT response for anxiety and obsessive-compulsive disorders. We hypothesized that pre-treatment as well as pre- to post-treatment changes in functional connectivity would be associated with improvement during CBT in a transdiagnostic sample.

**Methods:** Twenty-seven patients with primary social anxiety disorder (*n*=14) and primary body dysmorphic disorder (*n*=13) were scanned before and after 12 sessions of CBT targeting their primary disorder. Eligibility was based on elevated trait SFA scores on the Public Self-Consciousness Scale. Seed-based resting state functional connectivity associated with symptom improvement was computed using a seed in the posterior cingulate cortex/precuneus that delineated a self-other functional network.

**Results:** At pre-treatment, stronger positive connectivity of the seed with the cerebellum, insula, middle occipital gyrus, postcentral gyrus, and precuneus/superior parietal lobule, and stronger negative connectivity with the putamen, were associated with greater clinical improvement. Between pre- to post-treatment, greater anticorrelation between the seed and precuneus/superior parietal lobule was associated with clinical improvement, although this did not survive thresholding.

**Conclusions:** Pre-treatment functional connectivity between regions involved in attentional salience, self-generated thoughts, and external attention predicted greater CBT response. Behavioral and self-report measures of SFA did not contribute to predictions, thus highlighting the value of neuroimaging-based measures of SFA.

**Clinical Trials Registration:** ClinicalTrials.gov Identifier: NCT02808702 https://clinicaltrials.gov/ct2/show/NCT02808702

## Introduction

Anxiety and obsessive-compulsive (OC) related disorders are the most prevalent class of psychiatric disorders in the U.S. and globally (Kessler et al., 2005). They are associated with significant economic and health-related burden (Global Burden of Disease Collaborative Network, 2019; Rehm & Shield, 2019). Cognitive behavioral therapy (CBT), widely considered the first-line psychological treatment for these disorders, targets core dysfunctional cognitions and avoided situations, which negatively reinforce anxiety (Harrison et al., 2016; Kindred et al., 2022). However, only one-third to one-half of individuals who receive CBT for these disorders experience clinical remission (Fernández de la Cruz et al., 2021; Springer et al., 2018), which emphasizes the need to better understand predictors and mechanisms of CBT non-response. More research on factors driving therapeutic change may make existing treatments more potent and reduce costs associated with pursuing unhelpful treatments.

No single behavioral, clinical, or demographic variable has emerged as a reliable pre-treatment predictor of treatment response for anxiety disorders (Erceg-Hurn et al., 2023; Schneider et al., 2015), which highlights the potential value of brain-based markers as predictors of response. Functional Magnetic Resonance Imaging (fMRI) measures have been increasingly integrated into CBT studies with the recognition that they may explain more variance in treatment response, compared to available measures. For example, studies employing task-related fMRI have shown that certain brain regions implicated in threat processing and emotion regulation predicted treatment response (Lueken et al., 2016; Pico-Perez et al., 2023). However, given tremendous heterogeneity in task paradigms studied in anxiety and OC disorders, which leads to translational challenges across disorders, multivariate analyses of brain connectivity may offer a better understanding of interactions between regions that allow for integrated function. In support, studies have demonstrated that multivariate functional connectivity (FC) measures at pre-treatment significantly predicted therapeutic response and outperformed symptom severity measures (Moody et al., 2021; Reggente et al., 2018; Whitfield-Gabrieli et al., 2016). From this emerging literature, anxiety and OC disorders may be better conceptualized as disorders of disrupted brain connectivity, rather than deficits in regional brain activation of threat processing structures of the brain. Thus, in this study, we examined predictors and correlates of CBT using resting state FC.

Maladaptive self-focused attention (SFA) is a candidate transdiagnostic cognitive mechanism, which may represent a promising biomarker of CBT response. SFA is a disposition to focus excessively on internally-generated thoughts, feelings, beliefs, and physical states (Ingram, 1990), which may be linked to treatment response. Despite its role in cognitive theories of social anxiety disorder (Clark & Wells, 1995; Rapee & Heimberg, 1997; Norton & Abbott, 2016; Spurr & Stopa, 2002) and body dysmorphic disorder (Veale, 2004), the majority of studies on SFA have been conducted in depression and little is understood about its transdiagnostic versus disorder-specific effects. In social anxiety disorder (SAD), SFA is theorized to contribute to seeing oneself from an observer’s perspective in social situations (Clark & Wells, 1995) and has been linked to increased state anxiety (Woody & Rodriguez, 2000). In one study, high (compared to low) socially anxious individuals displayed hyperactivation of the medial prefrontal cortex, temporoparietal junction, and temporal pole, when engaging inward versus outward attention (Boehme et al., 2015). These regions comprise anatomical components of the default mode network (DMN), which is a complex network involved in internally-generated cognition. Although aberrant DMN resting state FC has been implicated in SAD (Liao et al., 2010; Liu et al., 2015), no studies have directly linked DMN connectivity with SFA and studies have not directly examined SFA in body dysmorphic disorder (BDD). Despite being classified in separate DSM-5 categories, SAD and BDD share strong conceptual, phenomenological, and treatment overlap (Fang & Hofmann, 2010; Fang et al., 2013). Previously, we found that maladaptive SFA may be associated with aberrant FC between regions comprising the DMN and dorsal attention network (DAN) in patients with SAD and BDD, which may reflect dysfunction in shifting between internal and external attention and represent a neural signature of SFA (Fang et al., 2022).

In this study, we extended these findings by investigating the brain connectivity correlates of SFA as a potential predictor of CBT response in SAD and BDD. Given previous work linking maladaptive SFA with DMN connectivity (Fang et al., 2022), we hypothesized that connectivity with the DMN would be more sensitive predictors of CBT response than behavioral or self-report measures in this transdiagnostic sample. We also hypothesized that pre-post changes in FC related to SFA would be associated with clinical improvement after CBT.

## Methods and Materials

### Participants

Participants were adult patients (between 18-45 years of age) with primary SAD or primary BDD, as described previously (Fang et al., 2022). Participants were recruited from outpatient psychiatry specialty clinics at Massachusetts General Hospital and from the community through print and online advertisements. In addition to meeting criteria for primary SAD or BDD (confirmed using the Structured Clinical Interview for DSM-5 (SCID-5; First et al., 2015)), as well as cutoff scores on symptom severity rating scales (≥50 on the Liebowitz Social Anxiety Scale (LSAS; Liebowitz, 1987) for those with primary SAD or ≥20 on the Yale-Brown Obsessive-Compulsive Scale Modified for BDD (BDD-YBOCS; Phillips et al., 1997) for those with primary BDD), participants also displayed scores ≥18 (1SD above the normative mean) on the Public Self-Consciousness Scale-Revised (SCS-R; Scheier & Carver, 1985) reflecting elevated self-reported trait SFA. The Public SCS-R was chosen as the best measure of SFA in this population, as public forms of SFA may be specifically associated with anxiety, whereas private forms of SFA may be more linked to depression (Mor & Winquist, 2002). Example items include: *I care a lot about how I present myself to others, and I usually worry about making a good impression*. Exclusion criteria were as follows: (1) MR contraindications (e.g., claustrophobia, metal in body, pregnancy), (2) history of head injury, neurological disorder, or neurosurgery, (3) active suicidal or homicidal ideation, (4) lifetime manic, hypomanic, or psychotic episode, (5) past year alcohol or substance use disorder, (6) unstable dose of psychotropic medication or discontinuation of psychotropic medication <2 months prior to study baseline, and (7) current or past CBT (>10 sessions) or formal mindfulness/meditation training. Analyses are reported on a final sample of 27 treatment completers (77.78% female, *n*=21,) with a mean age of 27.29 years (SD = 4.43). Pre-treatment characteristics in each group, as well as reasons for ineligibility and attrition, are described in *Supplementary Table S1*.

### Self-Other Task

Participants completed the Self-Other task (Kelley et al., 2002) while undergoing fMRI during each scan. The Self-Other task is a widely-used event-related fMRI task that asks participants to make judgments about trait adjectives comprising one of three types: Self (“Does this adjective describe you?”), Other (“Does this adjective describe former U.S. President Barack Obama?”), and Uppercase (“Is this adjective printed in uppercase letters?”). Each trial lasted 2500 ms and consisted of a “cue” word (Self, Obama, Uppercase) above a central fixation and a unique trait adjective (“POLITE”) below a central fixation presented for 2000 ms. A total of 270 unique adjectives were selected from a pool of normalized adjectives (Anderson, 1968). Lists were counterbalanced for word length, number of syllables, and valence. Longer reaction times for self versus other indicated greater SFA and represented a behavioral measure of SFA.

### Treatment

Participants received 12 50-minute weekly individual sessions of CBT provided by the same clinician (A.F.). The Hofmann & Otto (2008) CBT for SAD and Wilhelm et al. (2013) CBT for BDD treatment manuals were implemented for patients with primary SAD or BDD, respectively. Both treatments rely on core CBT principles and followed identical treatment components: psychoeducation (sessions 1-2), cognitive restructuring (sessions 3-4), introduction of exposure and in-vivo exposure practice (sessions 5-11), and relapse prevention (session 12). The therapist completed the Patient Exposure and Response Prevention Adherence Scale (PEAS; Simpson et al., 2010) to assess homework compliance between sessions 7-12. Treatment response was defined by a standardized continuous variable based on percent symptom reduction between pre- to post-treatment on the respective interviewer-rated symptom severity scale (LSAS for primary SAD or BDD-YBOCS for primary BDD).

### Procedure

The initial evaluation involved a diagnostic interview using the SCID-5, clinician-administered symptom assessments (e.g., LSAS to assess SAD severity or BDD-YBOCS to assess BDD severity, Brown Assessment of Beliefs Scale (BABS; Eisen et al., 1998) to assess levels of insight, and Clinical Global Impressions-Severity (CGI-S; Guy, 1976) to assess clinical severity), and self-report measures of depression severity using the Beck Depression Inventory-II (BDI-II; Beck et al., 1996), rumination using the Ruminative Responses Scale (RRS; Treynor et al., 2003), handedness using the Edinburgh Handedness Inventory (EHI; Oldfield, 1971), and estimated verbal IQ using the Wide Range Achievement Test-4 (WRAT-4; Wilkinson & Robertson, 2006). Clinician-rated assessments were conducted by the same independent evaluator (R.J.J.) at pre-treatment, immediately after session 6 (mid-treatment), and immediately after session 12 (post-treatment). Participants returned for a pre-treatment MRI scan 7-10 days of the initial evaluation before beginning CBT, and again for a post-treatment scan 7-10 days after the final treatment session. The study was approved by the Partners Human Research Committee and all participants provided written informed consent.

### MRI Acquisition

Participants were scanned in a 3T Siemens Prisma scanner (Siemens Medical Systems, Iselin, NJ) equipped for echo planar imaging and a 64-channel head coil. Except for 1 patient with BDD, all participants who had never undergone an MRI scan completed a mock MRI scan to acclimate to the scanning environment and train to remain still. A high-resolution anatomical scan was acquired using a T1-weighted 3D multiecho magnetization-prepared rf-spoiled rapid gradient-echo MEMPRAGE sequence with EPI based volumetric navigators for prospective motion correction and selective reacquisition (Tisdall et al., 2012) (TR=2530ms, flip angle=7°, TEs=1.7ms/3.6ms/5.4ms/7.3ms, iPAT=2, FOV=256mm, 176 in-plane sagittal slices; voxel size=1mm^3^ isotropic, scan duration 5m 34s). Two resting state scans were acquired with a T2*-weighted gradient echo sequence for blood oxygen level dependent (BOLD) contrast (TR=2000ms, flip angle=85°, TE=30ms, FOV=205mm, 32 continuous horizontal slices parallel to the intercommissural plane, voxel size=3.2x3.2x3.3mm, interleaved, scan duration 6 min 6s). The resting state sequences included prospective acquisition correction (PACE) for head motion to adjust slice position and orientation during image acquisition (Thesen et al., 2000). Participants were instructed to keep their eyes open for the duration of the scan.

### MRI Preprocessing and Data Quality

Preprocessing steps were performed using the CONN toolbox version 17 (Whitfield-Gabrieli & Nieto-Castanon, 2012), which employs routines from the Statistical Parametric Mapping software (SPM8; Wellcome Trust Centre for Neuroimaging, London, UK). Four initial volumes of each resting state run were discarded to allow for T1 equilibration. Images were segmented into gray matter, white matter, and cerebrospinal fluid masks, corrected for slice timing, and spatially realigned to the reference image. The volumes were then normalized to the MNI template provided in SPM8, resampled to 2mm voxels, and spatially smoothed using an 8mm full width at half maximum Gaussian kernel. A temporal band-pass filter of .008-.09 Hz was applied to the time series. Nuisance variables (white matter, cerebrospinal fluid, movement parameters) were addressed using the anatomical CompCor method (Behzadi et al., 2007; Chai et al., 2012). CONN uses the Artifact Detection Tools (ART; http://www.nitrc.org/projects/artifact_detect/) to identify artifactual volumes with >0.9 mm head displacement in the x, y or z directions or global mean intensity more than 5 SDs above the entire scan. Residual head motion (after PACE) was calculated using Power et al.’s (2012) measure of Framewise Displacement that takes into account both translational and rotational instantaneous motion.

### Treatment Response Analyses

To examine pre-post changes in SAD and BDD symptoms, delusionality, depression severity, and trait maladaptive SFA, we conducted linear mixed models using restricted maximum likelihood, a compound symmetry variance-covariance structure, and with time as a repeated random effect.

### Functional Connectivity Analyses

As previous research showed that trait SFA was linked to the posterior cingulate cortex (PCC) node of the DMN (Fang et al., 2022), we first conducted our analyses using the same seed region (in the PCC) as our previous analysis. However, results were stronger when restricting our analysis to a seed region that overlapped with the original seed, but was more specific to self-other processing based on meta-analytic evidence (Murray et al., 2012; 2015). We therefore selected this more specific ventral region of the PCC, as evidence also suggests that the ventral PCC may form a core self system within the DMN (Davey et al., 2016). We therefore drew a 10mm sphere around the peak coordinate of the PCC/precuneus [MNI: 2 -61 26], which was differentially responsive to self-other-processing (Murray et al. 2012; 2015). Seed-to-voxel FC analyses were conducted to assess FC associated with treatment response scores for treatment completers. First level FC maps were generated by extracting the average BOLD time series from the seed and correlating it with every gray matter voxel in the whole brain. Residual head motion parameters (rotations in x, y, and z directions and their first-order temporal derivatives) and artifactual volumes (flagged by Artifact Detection Tools) were scrubbed in CONN. Correlation values were then transformed to Fisher’s z to yield a map for each resting state run in which the value at each voxel represented connectivity with the seed. Two resting state runs for each scan (pre-treatment and post-treatment) were averaged.

For the prediction analysis, we examined relations between treatment response (based on the transdiagnostic continuous measure of percent symptom reduction described in the Treatment section) and FC of the self-other processing network at baseline. Given our interest in FC related to trait SFA, connectivity with any regions showing significant associations with symptom reduction was extracted for individual participants and correlated with baseline Public SCS-R scores. For the treatment correlates analysis, we examined the relationship between pre-post changes in FC of the self-other network and treatment response. For both regression models, whole brain correction for multiple comparisons was applied using a voxel level uncorrected threshold of p < 0.005 and a false discovery rate (FDR)-corrected cluster threshold of *p* < 0.05. Baseline symptom severity did not predict CBT response and was therefore not included in these models.

### Integration Analyses

We compared a linear regression model with only self-report (Public SCS-R scores) and behavioral measures (reaction time differences between self versus other trials) of trait SFA, to a model that additionally included pre-treatment FC measures of trait SFA to predict percent symptom reduction, and evaluated the models using the area under the receiver operating characteristic curve (AUC) based on a ≥60% symptom reduction threshold for categorically defining response/non-response. This ≥60% threshold was selected to generate more balanced groups.

## Results

### Treatment Response

After CBT, patients showed significant reductions in symptom severity, delusionality/poor insight, depression severity, and trait maladaptive SFA. Overall treatment adherence measured by the PEAS was good (mean across sessions 7-12 = 5.05 ± 1.69). Pre-post changes in clinical measures are reported in *Supplementary Table S2*.

Mean percent symptom reduction during CBT was 53.70% ± 15.78 across both groups. Using a ≥ 60% symptom reduction categorical definition of treatment response, 6 patients with BDD and 9 patients with SAD did not meet responder criteria. There was no difference between SAD versus BDD patients in percent symptom reduction achieved (*t*(25)=1.03, *p* = 0.20), nor in the number of patients who achieved response (X^2^ = 0.90, *p* = 0.34).

### Pre-Treatment Predictors of CBT Response

Pre-treatment resting state FC of the seed with the cerebellum, insula, middle occipital gyrus, postcentral gyrus, and precuneus/superior parietal lobule (SPL) predicted greater symptom reduction during CBT, whereas connectivity with the putamen predicted less symptom reduction during CBT (Table 1; Figure 1). There was a significant correlation between extracted connectivity measures and Public SCS-R scores for only two regions: the precuneus/SPL (*r* = -0.43, *p* = 0.03) and the putamen (*r* = 0.45, *p* = 0.02). Greater baseline SFA was associated with less positive (more negative) FC between the PCC/precuneus and precuneus/SPL, and more positive (less negative) FC between the PCC/precuneus and putamen.

**Figure 1.**
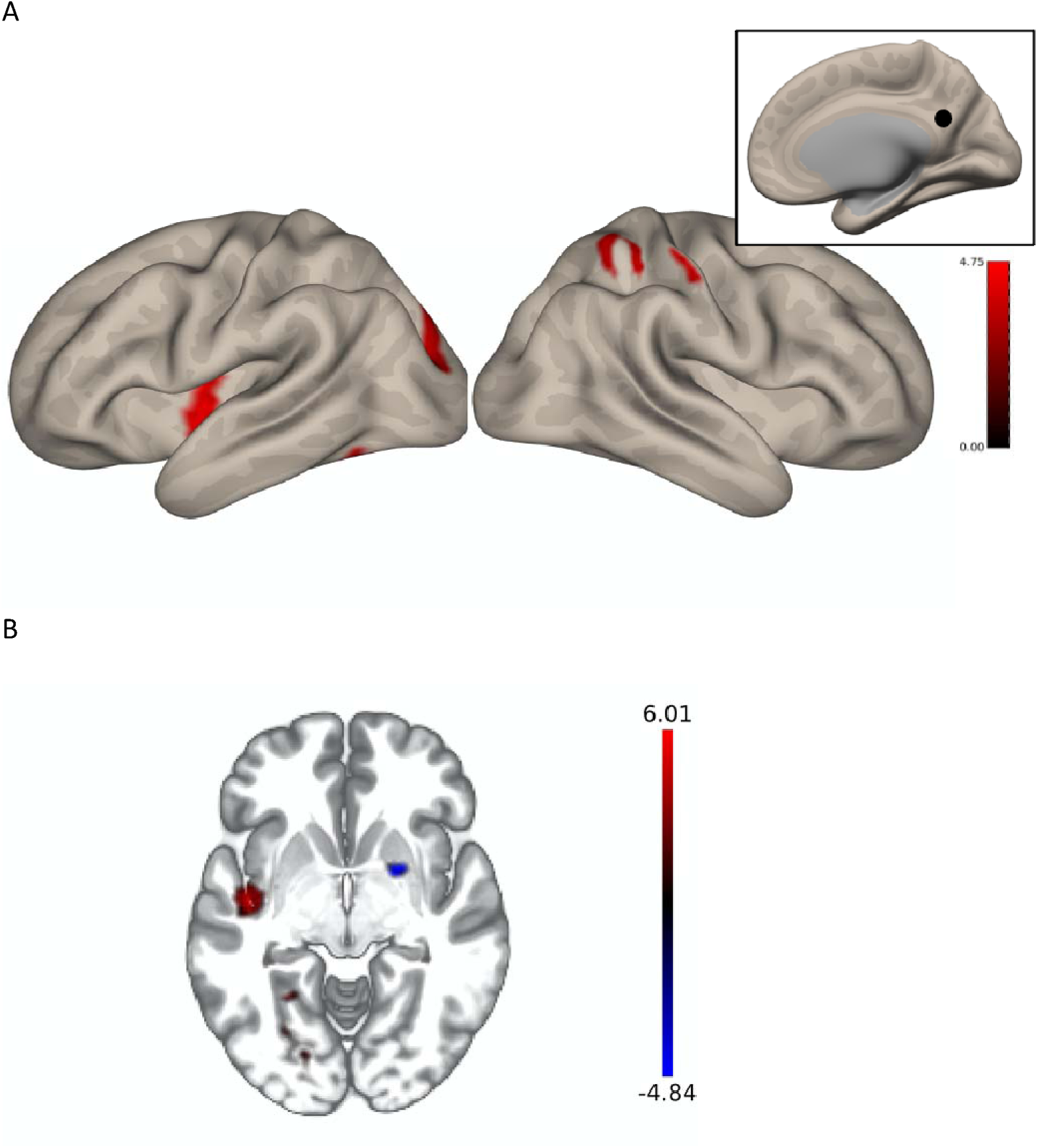
Pre-treatment functional connectivity associated with treatment outcome. Statistical map of correlation between connectivity and treatment outcome in (A) right lateral cortical surface (with an inset image depicting the seed location on the right medial surface) and (B) axial view at p_FDR_ < 0.05. Connectivity associated with greater symptom improvement in red and connectivity associated with less symptom improvement in blue. Color bars represent t-values.

**Table 1.** Clusters showing significant functional connectivity with self-other network seed which predict CBT response. All reported clusters are significant at *p*_FDR_ <0.05, two-sided, based on whole-brain correction. Montreal Neurological Institute (MNI) coordinates are given for peak voxel location.

| Region | Voxel<br>size | MNI Coordinates |  |  | T | BA |
| --- | --- | --- | --- | --- | --- | --- |
|  |  | x | y | z |  |  |
| *Pre-treatment connectivity predicting greater symptom improvement* |  |  |  |  |  |  |
| L cerebellum (culmen) | 794 | -30 | -66 | -24 | 6.01 | --- |
| L fusiform gyrus |  | -28 | -53 | -8 | 3.83 | 37 |
| L parahippocampus |  | -28 | -53.5 | -7.5 | 3.78 | 19 |
| L lingual gyrus |  | -22 | -78 | -2 | 3.78 | 18 |
| L middle temporal gyrus |  | -56 | -56 | -7.5 | 2.91 | 37 |
| L insula | 361 | -44 | -10 | -6 | 4.86 | 13 |
| L superior temporal gyrus |  | -46.5 | -8 | -5.5 | 4.40 | 22 |
| L claustrum |  | -40 | -10 | -5.5 | 4.25 | --- |
| L middle occipital gyrus | 306 | -24 | -88 | +16 | 3.90 | 19 |
| L cuneus |  | -2 | -80 | +16 | 2.85 | 17 |
| R postcentral gyrus | 292 | +62 | -20 | +50 | 4.54 | 2 |
| R inferior parietal lobule |  | +56 | -22 | +50 | 4.15 | 40 |
| R precuneus | 214 | +28 | -44 | +48 | 4.12 | 7 |
| R superior parietal lobule |  | +30 | -47.5 | +48 | 3.35 | 7 |
| R paracentral lobule |  | +26 | -40.5 | +48 | 2.92 | 5 |
| *Pre-treatment connectivity predicting less symptom improvement* |  |  |  |  |  |  |
| R putamen | 351 | +22 | +04 | -06 | -4.70 | --- |
| R lateral globus pallidus |  | +19 | +2 | -5 | -4.26 | --- |

### Associations Between Pre-Post Changes in Functional Connectivity and CBT Response

There were no regions surviving the voxel-level uncorrected threshold of *p* < 0.005 showing significant changes in FC that was associated with symptom reduction during CBT. However, at an uncorrected threshold of *p* < 0.01 (two-sided), change in FC with a region in the precuneus/SPL (BA7) extending into the postcentral gyrus (BA40) was associated with pre-post symptom improvement ([MNI: +38 -22 +28]; Figure 2A). We examined FC with the PCC/precuneus seed for patients separately at each timepoint based on a mask of the precuneus/SPL region identified in the pre-post analysis in order to evaluate the direction of change during treatment. CBT responders showed greater anticorrelations between the seed and precuneus/SPL over the course of treatment, whereas CBT non-responders showed greater positive correlations between these regions during treatment (*t*(25) = 4.16, *p* < 0.001) (Figure 2B). Change in FC was significantly associated with change in trait SFA based on Public SCS-R scores (*r* = 0.44, *p* = 0.02), such that as FC became more negative (anticorrelated) during CBT, SFA lessened.

**Figure 2.**
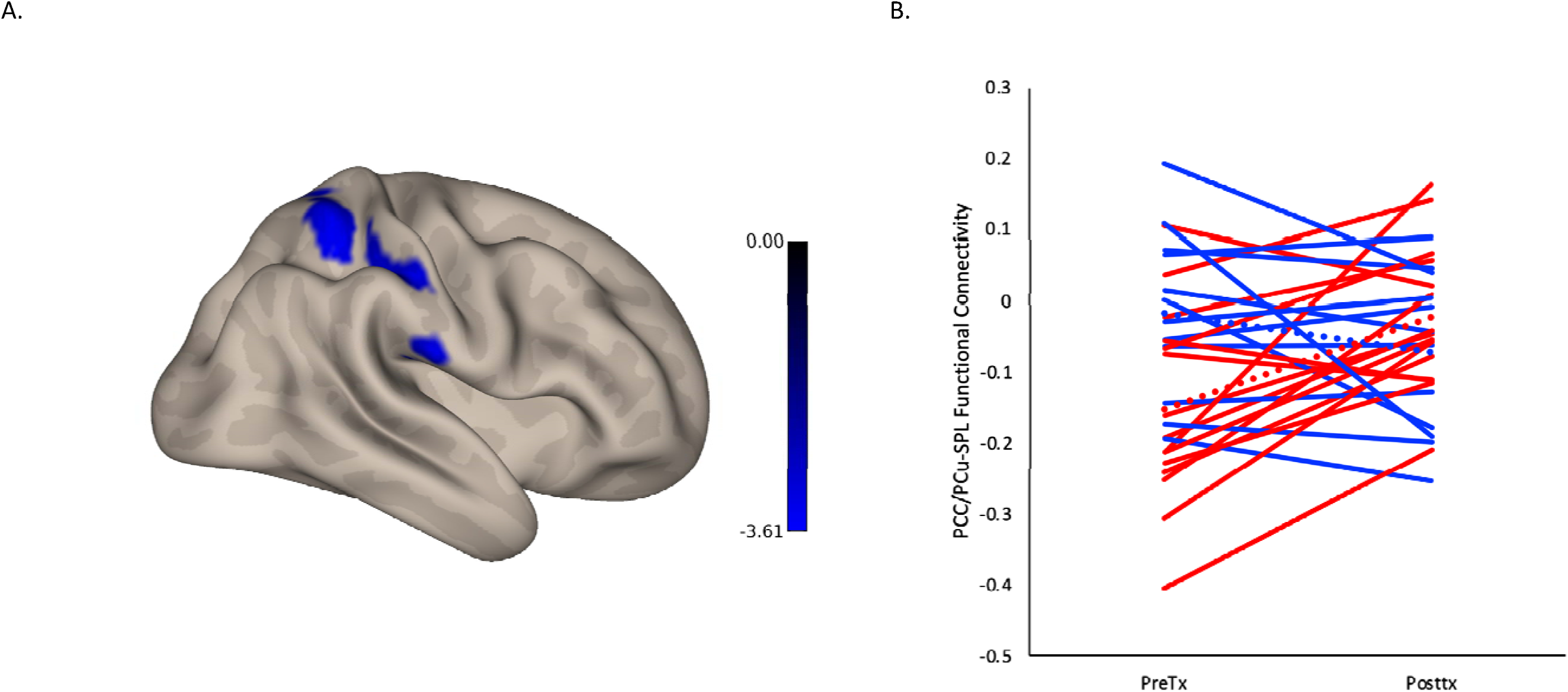
Changes in functional connectivity between pre- and post-treatment associated with the main effect of treatment. (A) Statistical map of correlation between connectivity and treatment effect at p_FDR_ < 0.05. (B) Individual trajectories of change in functional connectivity with precuneus/superior parietal lobule between pre- to post-treatment in treatment completers (*n*=27). Dotted lines reflect mean changes in connectivity. Those achieving ≥ 60% symptom improvement in blue, and those achieving < 60% symptom improvement in red. Color bar represents t-values.

### Integration Analyses

We examined FC between the PCC/precuneus seed and the precuneus/SPL for the integration analysis, as its association with SFA in this study replicated previous work (Fang et al., 2022). To avoid circularity, we examined FC with the PCC/precuneus seed for patients at the pre-treatment timepoint within a restricted mask from the pre-post analysis to be related to treatment response, rather than from the pre-treatment prediction analysis. Linear regressions showed that Public SCS-R scores and behavioral reaction time measures together only accounted for 1.7% of the variance in response, whereas pre-treatment FC with the precuneus/SPL, together with Public SCS-R scores and behavioral reaction time measures, accounted for 32.3% of the variance (Table 2). AUC for CBT responders was significant (*p* = 0.013, 95% CI: [0.60, 0.96]). As expected, in contrast to self-report and behavioral measures, which did not significantly predict symptom reduction after CBT, FC measures improved model predictions.

**Table 2.** Pre-treatment predictors of symptom improvement after CBT (*n*=27) Linear regression models comparing prediction of treatment response using only self-report and behavioral measures of SFA (Model 1) versus a combined model with self-report, behavioral, and neural measures (Model 2).

| Predictors | Model 1 |  |  | Model 2 |  |  |
| --- | --- | --- | --- | --- | --- | --- |
|  | *B* | 95% CI | *p* | *B* | 95% CI | *p* |
| Public SCS-R | -0.02 | [-0.07, 0.04] | 0.58 | 0.02 | [-0.04, 0.07] | 0.54 |
| Self-other reaction times | -0.12 | [-0.84, 0.60] | 0.74 | -0.10 | [-0.72, 0.51] | 0.74 |
| Connectivity with precuneus/superior parietal lobule |  |  |  | 0.66 | [0.24, 1.08] | 0.004 |
| $R^2$ | 0.02 | | | 0.32 | | |
| F for $R^2$ | 0.20 | | 0.82 | 3.65 | | 0.03 |

## Discussion

This is the first study to examine the neural correlates of SFA as a predictor and correlate of CBT response in any disorder, and the first to be done transdiagnostically. Our results suggest that CBT may be more beneficial for those with SAD or BDD who begin treatment with greater resting state FC with certain brain regions involved in self-referential and attentional processing, namely the cerebellum, insula, precuneus, and SPL.

Our finding that pre-treatment FC with the precuneus/SPL correlated with trait SFA is consistent with previous work on the inverse relationship between PCC-SPL connectivity and SFA (Fang et al., 2022). As the PCC is a functionally heterogeneous region thought to serve as a DMN hub, and the SPL is an anatomical component of the DAN, PCC-SPL connectivity may capture the internal and external attentional processes associated with SFA. This aligns with evidence that the DMN and DAN are intrinsically anticorrelated at rest to support internal and external foci of attention (Andrews-Hanna et al., 2014; Fox et al., 2005). Although the precuneus has also been identified as an important node of the DMN as an adjacent region to the PCC (Utevsky et al., 2014), the dorsal component of the PCC/precuneus has shown connectivity with both the DMN and attention networks at rest (Leech et al., 2011), suggesting it may play a unique role in allocating attention between internal and external attention states.

Our findings also demonstrate that pre-treatment FC with the insula predicts CBT response, which confirm previous findings implicating the bilateral insula as a region that distinguishes patients with maladaptive SFA from controls with low SFA (Fang et al., 2022). In that analysis, patients displayed decreased, whereas controls displayed increased, PCC connectivity with the insula. The anterior division of the insula is involved in interoceptive awareness (Menon & Uddin, 2010), and represents a key node of the salience network, which may regulate balance between external perception and internally-generated thoughts (Andrews-Hanna et al., 2014). Excessive focus on the physiological symptoms of anxiety reflects one aspect of maladaptive SFA (Ingram, 1990; Norton & Abbott, 2016). Previous work showed that neural activation in the anterior insula was positively correlated with trait SFA in high socially anxious participants (Boehme et al., 2015). Together, our findings suggest that pre-treatment resting state FC between default and attention networks may be predictors of CBT response.

A growing literature suggests involvement of the cerebellum in fear learning (Lange et al., 2015) and executive attention (Clark et al., 2022). FC studies in SAD and PTSD have reported pre-treatment connectivity with the cerebellum that predicted response to CBT and prolonged exposure therapy, respectively (Klumpp et al., 2014; Simmons et al., 2013). Furthermore, frontostriatal abnormalities have been more commonly reported in OCD and BDD (Feusner et al., 2010), but have also been found in SAD (Anteraper et al., 2014). Our findings indicate that pre-treatment PCC/precuneus-putamen connectivity was positively correlated with trait SFA and may be particularly predictive of CBT non-response. Early identification of CBT non-responders may generate hypotheses regarding mechanisms of non-response and lead to alternative therapies with greater success.

We found that a neuroimaging-based measure of maladaptive SFA (compared to self-report and behavioral measures) explained a greater proportion of variance in CBT response. Excessive focus on negative internal thoughts due to SFA may hinder one’s ability to integrate new learning during exposure (Clark & Wells, 1995), which may be critical for fostering inhibitory associations that compete with the strength of original fear associations (Craske et al., 2014). Thus, successful CBT outcomes may require some minimum attentional resources to optimize learning. Indeed, experimental manipulations of SFA have been associated with greater anxiety (Leigh et al., 2021; Woody & Rodriguez, 2000) and task interference (Judah et al., 2016), and attending to the external environment (away from internal self-focus) may enhance the effects of exposure (Wells & Papageorgiou, 1998).

Pre-post changes in FC showed remarkable overlap with regions identified in the pre-treatment data suggesting that FC with the precuneus/SPL is malleable and that this connectivity may become increasingly anticorrelated in CBT responders. Pre-post changes in FC also correlated with changes in trait SFA, suggesting that SFA itself may also be malleable during treatment. Improvements in SFA may be associated with a reduced reliance on negative internal information and greater external awareness. This is consistent with evidence that CBT leads to acute reductions in SFA during social interactions in patients with SAD (Hofmann, 2000; Kampmann et al., 2019; Woody et al., 1997), and that improvements in SFA may be an important mechanism of change in CBT (Hedman et al., 2013).

Clinically, what is striking about FC with the precuneus/SPL emerging in both the prediction and treatment correlates results is that some individuals with this specific pattern of lower FC at pre-treatment were still able to fully benefit from CBT and achieve full response by post-treatment, suggesting that having more information beyond pre-treatment could potentially change treatment recommendations. Future research should examine the potential utility of repeated scans during CBT to assess treatment progress (e.g., to assess whether attentional networks are moving in the expected direction) and inform decisions to continue CBT or refer for alternative treatment, such as with attentional training interventions.

Our study had limitations. First, our study design allowed for only correlational inferences. Future work may evaluate FC differences due to experimentally-induced SFA (e.g., through false heartrate feedback) to provide causal evidence in support of network interactions responsive to internal versus external attention manipulations. Second, we did not employ a treatment control or waitlist condition to address the possibility that FC effects were due to CBT, or the mere passage of time, respectively. Despite this, we showed that patients adhered to the protocol and practiced between-session exposures (based on the PEAS), which supports the notion that symptom improvement could be attributed to skills introduced during CBT. Finally, analyses were conducted on individuals who completed treatment and do not address potential confounds associated with discontinuing treatment. In summary, for self-focused individuals with SAD and BDD, neuroimaging-based measures of SFA may be more sensitive predictors of CBT response, compared to behavioral or self-report measures. Having more information about FC changes during CBT may offer more meaningful treatment recommendations than pre-treatment data alone.

## Data Availability

All data produced in the present study are available upon reasonable request to the authors.

## Acknowledgments

The authors would like to acknowledge Dr. Susanne Hoeppner for database management, as well as Rachel Porth, Jin Shin, and Lucas Petre for assistance with data collection.

## Declarations of Interest

J.D.F. receives consultation fees from NOCD, Inc.

R.J.J. has received salary and research support from the International OCD Foundation, the Charles A. King Trust Postdoctoral Research Fellowship Program (Bank of America, N.A., Co-Trustees), the Harvard University Mind Brain Behavior Initiative, the Harvard University Department of Psychiatry Livingston Fellowship Award, and the Harvard University Pershing Square Fund for Research on the Foundations of Human Behavior, and the National Institute of Mental Health (NIMH; K23 MH120351). She is also paid as a faculty member of the Massachusetts General Hospital Psychiatry Academy for filming role plays for course content and for moderating the course discussion boards. She receives book royalties from Hogrefe Publishing.

S.W. is a presenter for the Massachusetts General Hospital Psychiatry Academy in educational programs supported through independent medical education grants from pharmaceutical companies. She has received royalties from Guilford Publications, New Harbinger Publications, Springer, and Oxford University Press. Dr. Wilhelm has also received speaking honoraria from various academic institutions and foundations, including the International Obsessive Compulsive Disorder Foundation, the Tourette Association of America, and the Centers for Disease Control and Prevention. In addition, she received honoraria for her role on the Scientific Advisory Board for One-Mind (PsyberGuide), Koa Health, Inc, and Noom, Inc. Dr. Wilhelm has received research and salary support from Koa Health, Inc.

No other co-authors have any declarations of interest.

## Contributors

Angela Fang: Conceptualization, Methodology, Formal analysis, Investigation, Data curation, Writing – Original Draft, Writing – Review and Editing, Project administration, Funding acquisition; Bengi Baran: Formal analysis, Data curation, Writing – Review and Editing; Jamie Feusner: Formal analysis, Writing – Review and Editing; Luan Phan: Formal analysis, Writing – Review and Editing; Clare Beatty: Data curation, Project administration, Writing – Review and Editing; Jessica Crane: Data curation, Writing – Review and Editing; Ryan Jacoby: Investigation, Writing – Review and Editing; Dara Manoach: Conceptualization, Methodology, Resources, Supervision, Writing – Review and Editing, Funding acquisition; Sabine Wilhelm: Conceptualization, Methodology, Formal analysis, Resources, Supervision, Writing – Review and Editing, Funding acquisition.

## Supplementary Material

### Participant Characteristics

Fifty total participants consented to the study, of which 30 were eligible and began treatment. Reasons for ineligibility included: did not have primary SAD or BDD (n=9), did not meet cutoff score on the Public SCS-R (n=4), concurrent or past exclusionary therapy or meditation (n=4), and planning pregnancy (n=1). Two additional participants were deemed eligible but withdrew prior to starting treatment due to the time commitment. Of the 30 participants who began treatment, 3 participants withdrew, all right after the third treatment session, for varying reasons (moving out of state (n=1), pursuing more intensive treatment out of state (n=1), and being lost to follow-up (n=1)). Thus, the overall attrition rate was 10% (3/30).

Individuals with SAD (n=14) were older than those with BDD (n=13) and there were differences in self-identified racial group membership (Table S1), but there were no other differences between those with SAD versus BDD in sex, ethnicity, highest level of education, handedness, and estimated verbal IQ. At study baseline, two patients with BDD were taking citalopram, another was taking amphetamine, and one patient with SAD was taking escitalopram. All medications were taken at a stable dose for at least two months prior to study enrollment. Six participants met criteria for current major depressive disorder (n=3 with primary BDD, n=3 with primary SAD).

---

**Table S1.** Sample characteristics by diagnostic group.

| Characteristic | BDD (n=13) | SAD (n=14) | *t/X* | *p* |
| --- | --- | --- | --- | --- |
|  | Mean ± SD<br>/ *n* (%) | Mean ± SD<br>/ *n* (%) |  |  |
| Age (years) | 23.85 ± 2.27 | 27.29 ± 4.43 | -2.51 | 0.02 |
| Sex (female) | 9 (69.23) | 12 (85.71) | 1.06 | 0.30 |
| Race |  |  | 9.20 | 0.03 |
| Black/African American | 2 (15.38) | 0 |  |  |
| Asian | 0 | 6 (42.86) |  |  |
| More than one race | 1 (7.69) | 0 |  |  |
| Caucasian | 10 (76.92) | 8 (57.14) |  |  |
| Ethnicity (Hispanic) | 3 (23.08) | 1 (7.14) | 1.36 | 0.24 |
| Education |  |  | 2.03 | 0.57 |
| Some college | 1 (7.69) | 1 (7.14) |  |  |
| College graduate | 8 (61.5) | 7 (50.00) |  |  |
| Some post graduate | 1 (7.69) | 0 |  |  |
| Post graduate degree | 3 (23.08) | 6 (42.86) |  |  |
| Handedness | 77.69 ± 20.17 | 76.43 ± 44.35 | 0.09 | 0.93 |
| WRAT-4 | 72.23 ± 24.17 | 75.57 ± 18.08 | -0.41 | 0.69 |
| Head motion<sup>a</sup> | 0.20 ± 0.13 | 0.14 ± 0.05 | 1.57 | 0.14 |
| Number of artifactual volumes<sup>a</sup> | 12.54 ± 13.41 | 8.71 ± 2.81 | 1.04 | 0.31 |
| BDI-II | 14.62 ± 11.64 | 15.50 ± 13.96 | -0.18 | 0.86 |
| LSAS | --- | 78.64 ± 15.32 |  |  |
| BDD-YBOCS | 29.31 ± 6.30 | --- |  |  |
| BABS | 14.85 ± 4.14 | 14.07 ± 2.81 | 0.57 | 0.57 |
| Public SCS-R | 19.92 ± 1.12 | 20.00 ± 1.30 | -0.16 | 0.87 |
| RRS | 50.85 ± 12.36 | 51.79 ± 15.87 | -0.17 | 0.87 |

*Notes.* BDI-II = Beck Depression Inventory-II. LSAS = Liebowitz Social Anxiety Scale. BDD-YBOCS = Yale-Brown Obsessive-Compulsive Scale Modified for BDD. BABS = Brown Assessment of Beliefs Scale. Public SCS-R = Public Subscale of Self-Consciousness Scale-Revised. RRS = Ruminative Responses Scale.
<sup>a</sup>Averaged across both pre-treatment resting state scans.

**Table S2.** Pre-post changes during CBT for patients (*n*=27)

| Characteristic | Pre-Treatment |  | Post-Treatment |  | *F* | *p* |
| --- | --- | --- | --- | --- | --- | --- |
|  | M ± SD | 95% CI | M ± SD | 95% CI |  |  |
| LSAS | 80.27 ± 3.47 | [73.12, 87.42] | 39.06 ± 3.58 | [31.69, 46.43] | 96.60 | <0.001 |
| BDD-YBOCS | 30.20 ± 1.81 | [26.39, 34.01] | 13.50 ± 1.87 | [9.59, 17.42] | 141.35 | <0.001 |
| BABS | 14.57 ± 0.78 | [12.99, 16.14] | 6.65 ± 0.81 | [5.01, 8.29] | 91.37 | <0.001 |
| BDI-II | 16.27 ± 2.29 | [11.63, 20.90] | 10.09 ± 2.35 | [5.334, 14.85] | 10.98 | 0.003 |
| Public SCS-R | 20.07 ± 0.60 | [18.87, 21.26] | 15.99 ± 0.63 | [14.73, 17.24] | 28.61 | <0.001 |

## Notes

### Clinical Trial

NCT02808702

### Funding Statement

This study was funded by an NIMH Career Development Award (K23 MH109593) to A.F.

### Author Declarations

Ethics approval for the current study protocol was obtained by the Partners Human Research Committee (Protocol#2016P000523).

